# Quality measures of two-stage newborn hearing screening: Systematic review with a Bayesian meta-analysis

**DOI:** 10.1101/2024.01.29.24301931

**Authors:** Kirsi Manz, Uta Nennstiel, Carola Marzi, Ulrich Mansmann, Inken Brockow

**Affiliations:** Institute for Medical Information Processing, Biometry and Epidemiology (IBE), Faculty of Medicine, LMU Munich, Munich, Germany; Bavarian Health and Food Safety Authority, Oberschleißheim, Germany

## Abstract

**Background:** Newborn Screening for hearing impairment (NHS) is a crucial public health issue worldwide. Often, a two-stage screening with two different testing approaches is used. We aimed to investigate the optimal screening algorithm, based on data from the literature published in the last 30 years. A particular focus of the study was to synthesize the existing evidence on two-stage newborn hearing screening regarding the refer rate (RFR), the percentage of children that did not pass the second test or were lost after the first test.

**Methods:** We searched MEDLINE and Scopus for studies on two-stage NHS using transitory evoked otoacoustic emissions (TEOAE) or automated auditory brainstem response (AABR). All studies on newborns who received their first test as an inpatient and a second test up to one month later were eligible. Random effects meta-analysis and Bayesian modeling were performed to estimate RFR as well as effects of the second test phase on the RFR. Risk of bias was assessed using QUADAS-II. The unfunded study was registered in PROSPERO (CRD42023403091).

**Results:** Eighty-five study protocols, including over 1,12 million newborns, met the inclusion criteria. Certainty in the evidence was rated as moderate. The RFR was higher when the test method was changed than without a change of method (AABR-AABR: RFR = 1.3% (95% confidence interval (CI): 0.9, 1.8%), TEOAE-TEOAE: RFR = 2.7% (CI: 2.2, 3.2%), TEOAE-AABR: RFR = 3.9% (CI: 2.9, 5.1%), AABR-TEOAE: 5.9% (CI: 5.0, 6.9%).

**Conclusions:** Strategies that did not involve changes to the screening method had lower RFR.

## Introduction

A properly functioning auditory system is essential for a child’s acquisition of spoken language. Early intervention is critical for age-appropriate spoken language development in children with hearing impairment [1–5]. Therefore, the Joint Committee on Infant Hearing strives for a start of interventions no later than at three months of age [6]. Since the late 1990s, two screening methods have been available which allow a very early diagnosis: the measurement of transitory evoked otoacoustic emissions (TEOAE) and automated auditory brainstem response (AABR) [7]. With these testing methods, a prevalence for permanent bilateral congenital hearing impairment of about 1,3 per 1,000 newborns in developed countries [8], good treatment options with hearing aids, cochlear implants and early intervention, and a positive cost-benefit balance [9–11] congenital hearing disorders are suitable as a target disease for newborn screening [12,13]. Universal newborn hearing screening (UNHS) was included in standard care in Germany in 2009, as specified by the Federal Joint Committee (G-BA) in §§ 47 to 57 of the Pediatrics Directive. The Pediatrics Directive outlines a two-step screening algorithm in well babies, beginning with initial TEOAE measurement in both ears, followed by bilateral AABR measurement if the initial test is not passed, and defines quality criteria. Distortion product otoacoustic emission (DPOAE) is not allowed as a screening method in this algorithm. DPOAE thresholds are limited to 50 dB HL, and the goal of newborn hearing screening (NHS) is to detect bilateral hearing loss with a threshold of 35 dB HL [14].

An important quality factor of a screening program is the refer rate (RFR). In the NHS, this refers to the percentage of screened infants who require referral to a pediatric audiologist for further diagnostic examinations after not passing the screening tests (positive screening). This includes newborns who are lost-to-follow-up after not passing the initial test, as well as those who receive a “fail” result on the second test. In the Pediatrics Directive the RFR should not exceed 4 %. False positive findings are of particular concern as they can cause anxiety for affected families and increase the demand for scarce resources in pediatric audiology practices and outpatient clinics.

Evaluations of the German NHS for the years 2011/12 [8] and 2017/18 [15,16] have shown that the recommended screening-algorithm is often not followed. In more than 50% of cases where infants do not pass the first test, a second TEOAE measurement is performed instead of the required AABR. Additionally, this second TEOAE test yielded a “fail” result in only about 10% of cases, compared to 20% for infants who underwent a second test using AABR. The failure rate of the second test was particularly high when the screening method was altered. Analysis of these data demonstrated that the second test showed the lowest failure rate with the TEOAE / TEOAE algorithm at 9.62%, while the highest rate was observed with TEOAE / AABR at 26.59%. Accordingly, international recommendations suggest performing a second TEOAE test after the initial TEOAE result was “fail” in newborns without risk factors for hearing impairment (“well-babies”) [6,17].

This study reviewed the current literature to investigate the quality of available screening tests and the optimal screening algorithm based on data published in the last 30 years. Specifically, the study focused on synthesizing the existing evidence related to RFR in the two-stage NHS to determine the best two-step screening algorithm based on one of the two testing devices (TEOAE and AABR).

## Materials and methods

### Model

The first step was to formalize the two-stage screening process on which our meta-analysis was based.

The study population consists of newborns with relevant hearing impairment (“diseased”, D^+^) and those without (“healthy”, D^-^). The prevalence of hearing impairment in the population is denoted by π. The first stage of screening is performed using a method with a sensitivity of SE1 and specificity of SP1. Therefore, the test positivity rate of the first stage (PR1), which includes true positives (TP1) and false positives (FP1), is given by: PR1 = TP1 + FP1 = π*SE1 + (1-π)*(1-SP1). These newborns are considered “*failed”* and should receive a second test. The rate of positive results from the first stage is also referred to as the “failure rate”.

The proportion of newborns whose first test result was “pass” and who therefore leave the screening process is the negative rate NR1 = π*(1-SE1) + (1-π)*SP1. However, not all newborns with a positive first test proceed to the second stage: The proportion ρ of positively tested newborns is *lost*. This loss is assumed to be independent of hearing status (newborns drop out for reasons unrelated to the first test result). Thus, a proportion of (1-ρ) newborns undergo the second stage screening test.

The proportion of SE2 is identified as positive among the true positives of the first test. Accounting for the loss, this yields TP2 = TP1*(1-ρ)*SE2 = π*SE1*(1-ρ)*SE2. Similarly, the proportion of (1-SP2) is falsely identified as positive among the false positives of the first test. Accounting for the loss, we obtain FP2 = FP1*(1-ρ)*(1-SP2) = (1-π)*(1-SP1)*(1-ρ)*(1- SP2).

The proportion of newborns with “fail” results in both tests, together with those who are lost after a first positive test, form the refer rate (RFR):

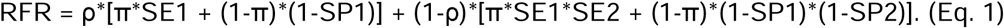

The structure of the two-stage model, together with the respective selection and loss to follow-up processes is shown in S1 Fig.

### Theoretical consideration of factors influencing the refer rate

We consider the relationship between the first stage positive rate (PR1 = π*SE1 + (1-π)*(1- SP1)) and the refer rate (RFR).

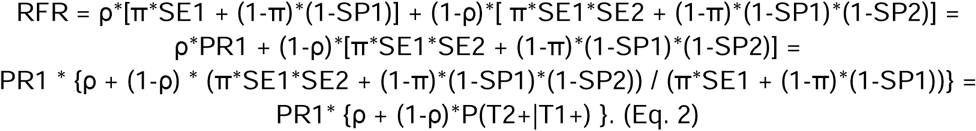

In case of a small prevalence π (close to 0, inserting π=0 in Eq.2), it holds

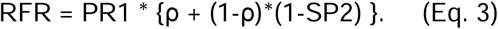

For small prevalence estimates, the RFR is linearly related to the failure (positive) rate of the first stage PR1. This linear relationship is determined by the loss rate ρ and the specificity of the second stage test SP2. Such a linear relationship between the RFR and failure rate can be exploited in a meta-regression. The quantity P(T2+|T1+) is called the conditional stage 2 positive rate (cPR2).

Equations 2 and 3 split the RFR into a component that quantifies the influence of the first stage and a component that quantifies the influence of the procedures after stage 1 (selection to stage 2 and quality of the second test). The RFR is the product of these two factors. It is of interest to quantify influence after stage 1. This will result from a meta-regression.

### Study protocol

The meta-analysis was prospectively registered in PROSPERO (CRD42023403091). The amended review protocol can be found at and downloaded from https://www.crd.york.ac.uk/prospero/display_record.php?ID=CRD42023403091.

### Eligibility criteria

For this review, the following PICO criteria were applied:

1. **(P)** Population: (Well) babies undergoing a two-stage hearing screening: (1) Initial screening as an inpatient in the maternity clinic; (2) Second test up to a maximum of one month later; (3) No use of the test method distortion product otoacoustic emissions (DPOAE); (4) Not exclusively newborns from NICU (neonatal intensive care unit).
2. **(I)** Intervention: Two-stage hearing screening using TEOAE, AABR, or a combination of both
3. **(C)** Comparator: not applicable
4. **(O)** Outcome: RFR after two screening steps

The population should preferably consist of well babies. Studies that included newborns with risk factors for hearing impairment or babies from the NICU were included if the study data did not clearly distinguish between well babies and these newborns. However, studies that included only newborns with risk factors or from the NICU were excluded from this review.

The review considered studies in which both the first and the second tests were completed within one month after birth.

### Search method

We searched MEDLINE using PubMed and Scopus for relevant articles without language or geographic restrictions from the time of their inception through February 9^th^, 2024. The following search strategy was used for both databases:

*(“newborn hearing screening”) OR (“neonatal hearing screening”) OR (“infant hearing screening”) OR (((“newborn screening”) OR (“neonatal screening”) OR (“infant screening”)) AND (“hearing”)) OR ((“hearing screening”) AND (“newborn” OR “neonatal” OR “infant”))*

A broader search strategy without specification of test method (TEOAE or AABR) was chosen to avoid missing relevant publications that may not specify the test method in the title or abstract. This decision was based on a small pilot search conducted to test the search strategy, which showed that including the type of screening test in the search resulted in missing some articles that were already known to be relevant to the review.

All articles had an abstract in English. If the full texts of the selected articles were not written in a language that the authors speak, online translation tools (DeepL, Google Translator) were used to translate them into English where possible. Only peer-reviewed publications were included.

### Study records

The search strategy was saved in Citavi version 6.11. The data from the selected publications were extracted into Excel spreadsheets.

One reviewer searched the information sources and screened the titles and abstracts of the identified studies for inclusion and classified each study as eligible or ineligible. The study was classified as potentially eligible if it could not be clearly excluded based on its title and abstract. The full texts of all (potentially) eligible studies were then retrieved and reviewed by two additional reviewers. Again, studies were marked as eligible or ineligible for inclusion and the selection was discussed with the first reviewer until all three reviewers agreed. The first reviewer extracted the required data and made the preliminary decision on study inclusion based on the availability of the data for extraction. The second reviewer double-checked the extracted data and made corrections, which were discussed with all three reviewers and led to the final inclusion of all studies with available data.

### Data extraction and items

Data extracted for the study description included the following: First author and year of publication, screening test combination (i.e., TEOAE-TEOAE, TEOAE-AABR, AABR- TEOAE, AABR-AABR), years of screening, country, number of newborns screened, name of screening device(s), time of first test and second test, and whether the study was conducted only in well babies or also in newborns with risk factors/ from the NICU.

Data extracted for quantitative analysis included the following: Number of newborns screened with the first test and type of first test (TEOAE/AABR), number of newborns who passed the first test, number of newborns who did not pass the first test, number of newborns who did not pass the first test and did not return for the second test (lost-to-follow-up), number of newborns screened for the second time and type of second test (TEOAE/AABR), number of newborns passing the second test, number of newborns who did not pass the second test, and number of newborns referred after not passing both screening tests.

Studies providing data on simultaneous TEOAE and AABR testing were added to both screening test combinations: TEOAE followed by AABR and AABR followed by TEOAE. Otherwise, such studies would have had to be excluded as the order of the tests was not sequential.

The following data were derived from the variables collected: The failure rate after the first test step was calculated as the quotient of the number of newborns who did not pass the first test and all newborns screened in the first step. Similarly, the failure rate after the second test step was calculated as the quotient of the number of newborns who did not pass the second test and all newborns screened in the second step. The RFR was calculated as the sum of the number of newborns who did not pass the second test and the number of newborns who did not pass the first test and did not return for the second test, divided by the total number of newborns screened.

### Outcomes and prioritization

Data were sought to calculate the RFR after the two-step NHS, with the failure rate after the first test included in the analysis. For the analysis, it was essential to obtain the number of infants who passed and did not pass each screening step. Therefore, studies that only reported the results after both screening steps could not be included in the review. This review focused mainly on well babies, but studies conducted in both well babies and newborns with risk factors or from the NICU were also included.

### Risk of bias in individual studies and publication bias

The quality of the included studies was assessed using a modified version of the QUADAS-II tool developed for diagnostic accuracy studies [18]. Of the original four domains, the “reference test” domain was not applicable to our study because we focused on the two-stage screening process without knowing the true hearing loss status. Therefore, we replaced the domain “index test” by “first test” and “reference test” by “second test”. The risk of bias for each included study was assessed independently by two reviewers.

Disagreements were resolved by discussion with a mediator. The risk of bias was assessed at the study level.

Publication bias was not expected to have a significant impact on the literature found. It was assumed that all studies on two-stage NHS would be worthy of publication, as they describe not only the quality of the NHS, but also its implementation and problems. Selective reporting within studies, e. g. favoring one of the two screening tests, is also unlikely to be a problem. Therefore, methods to assess the risk of publication bias were not used in this meta-analysis.

### Confidence in cumulative evidence

The strength of the overall body of evidence was assessed using Grading of Recommendations, Assessment, Development and Evaluation [19].

### Subgroup analyses

Subgroup analyses were not specified in the study protocol. However, based on the characteristics of the included studies, we decided to analyze only well-baby studies as a subgroup. An additional post hoc sensitivity analysis was performed to assess the effect of outliers in the TEOAE-TEOAE group.

### Statistical analysis

Continuous variables were summarized by median (minimum - maximum) or presented as box plots, and categorical variables were presented by frequency (%). Due to the exploratory nature of our study, adjustment for multiple testing was not considered. Statistical significance was claimed at 5% level (p<0.05) or for non-overlapping 95% credibility intervals. Random-effects meta-analysis of the RFR was performed using the R package *rmeta* and the DerSimonian-Laird approach. Heterogeneity indices Q and I^2 were calculated using the random effect estimates and random effect weights. For the visualization of the risk of bias, we used the source code of the *rob_summary* function of the *robvis* package to generate a similar graph adapted to our needs. Calculations were performed using WinBUGS version 1.4.3 [20] and R Version 4.3.2 [21]. All data and analysis scripts are available in the Open Science Framework repository at https://osf.io/nuk4p/.

## Results

The PRISMA flow diagram for the search and study selection process is shown in Fig 1. Out of the 5886 records identified (PubMed: n=3356, Scopus: n=2530), 2000 duplicates were removed prior to screening. From a total of 3886 records screened, a total of 3563 records were excluded because the titles and abstracts of these articles were not relevant to our research question. A full-text search was conducted on the remaining 323 records. Seven records could not be retrieved. Out of the 316 reports that were screened, 239 were excluded. In many cases this was due to a failure to provide the required data at each screening step (n=77), an unsuitable study design (n=50), or not performing the first or second test within the specified time frame of one month (n=44). Additional reasons for exclusion are listed in Fig 1. N=7 reports comprised more than one screening protocol (n=6 two protocols [22–27] and n=1 three protocols [28]). Of the reports with more than one protocol, three [25,27,28] provided data on simultaneous TEOAE and AABR testing and were included in both the TEOAE-AABR and AABR-TEOAE groups. A total of 77 reports with 85 study protocols were included in the meta-analysis.

**Fig 1.**
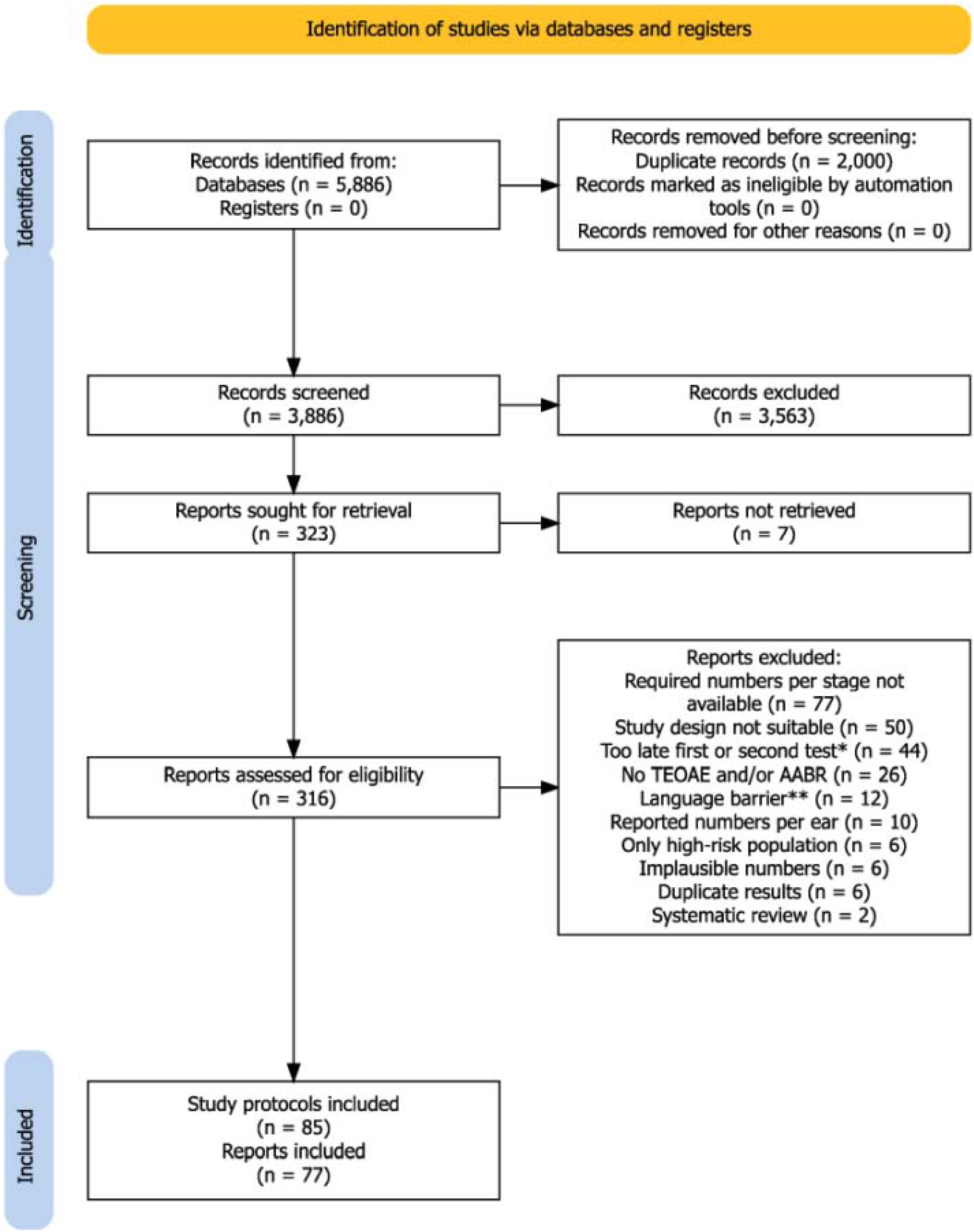
**PRISMA flow diagram of the search and study selection process.** *Includes also studies where the time point of the first or second test was not specified. **Language barrier refers to reports that could not be automatically translated into English online. A detailed list of the 239 excluded reports is available from the authors upon request.

The analysis included 85 study protocols from 77 reports (7 of those with more than one study protocol) with 1125617 newborns. Table 1 provides an overview of the study characteristics.

**Table 1:**
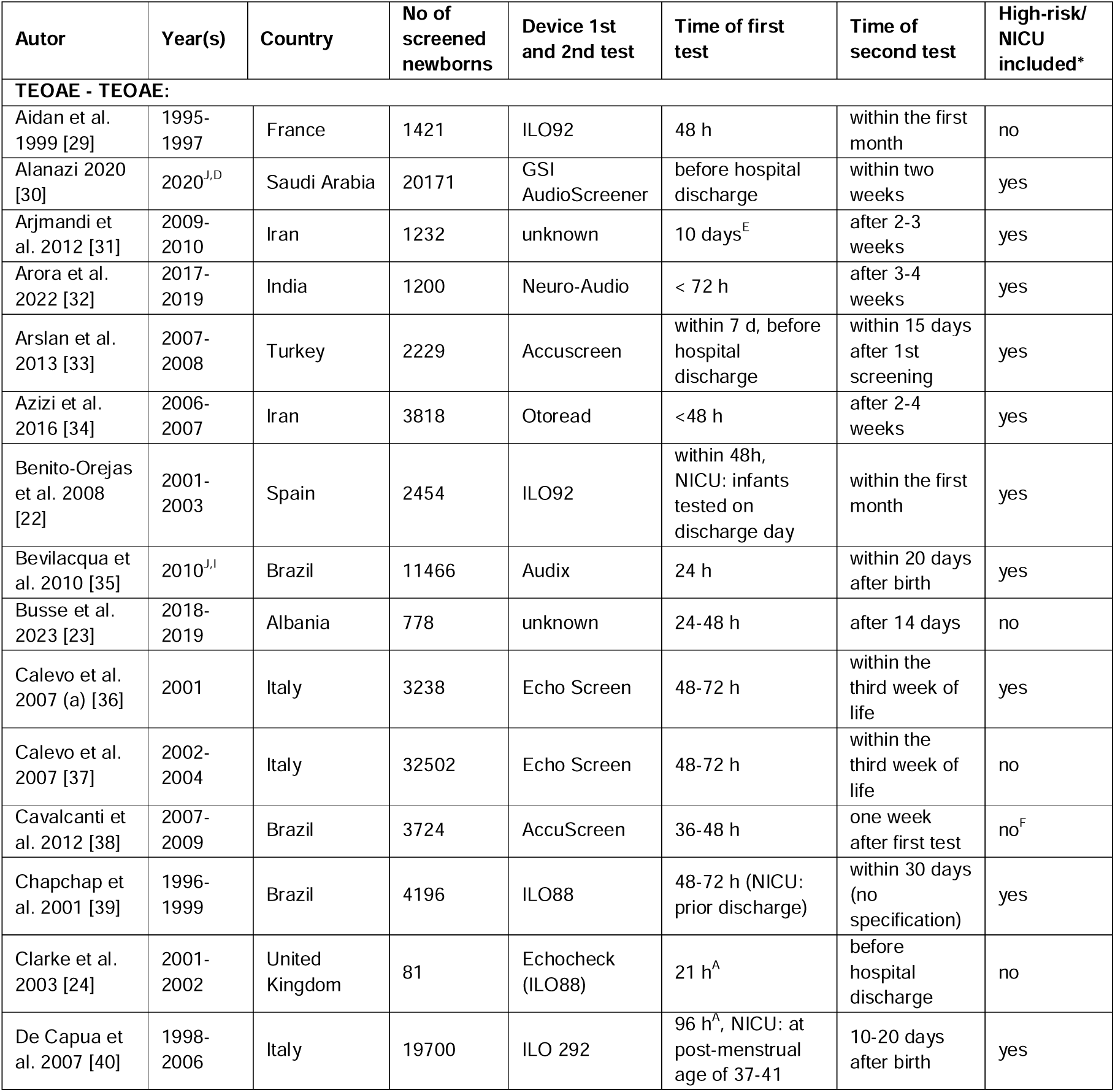
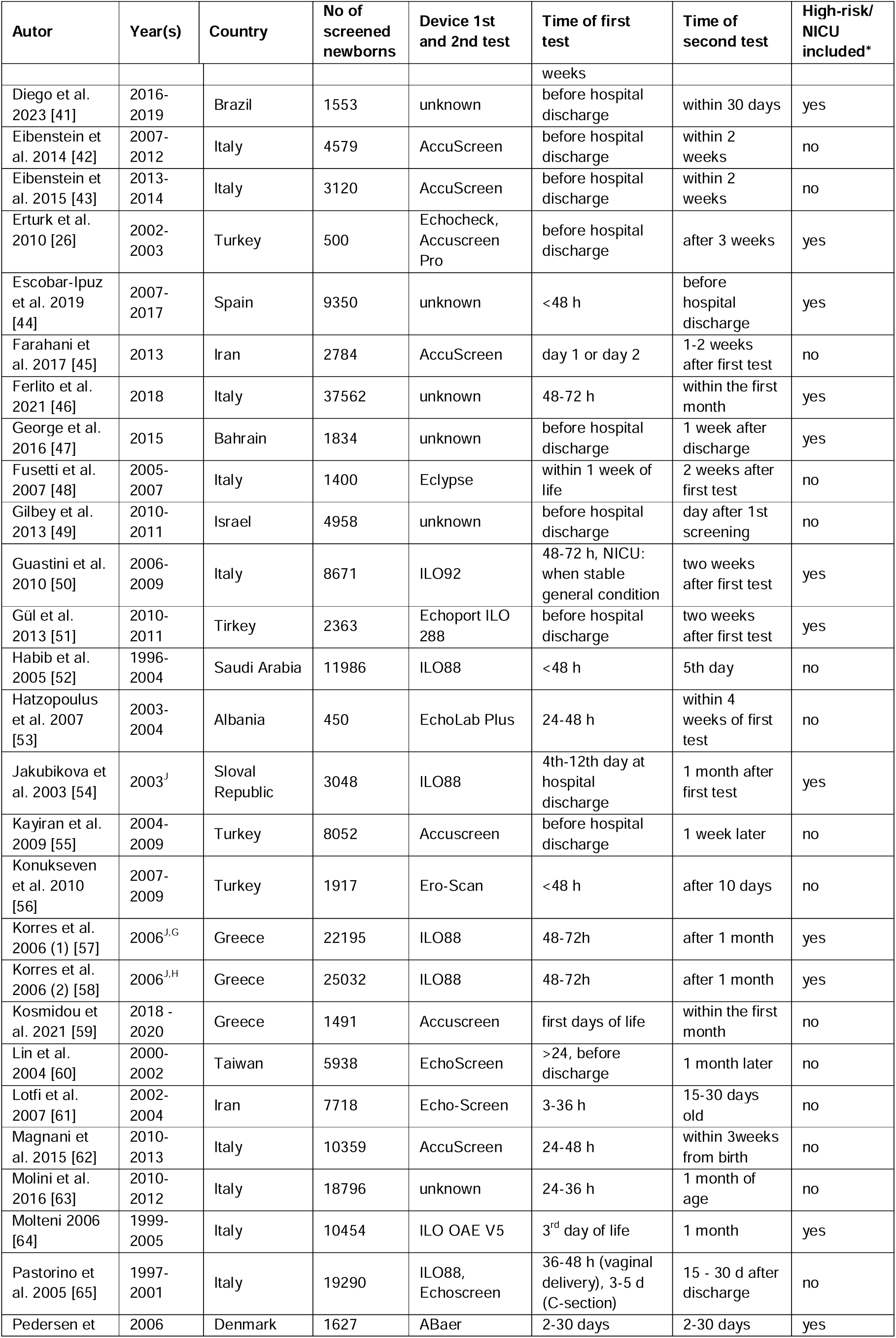
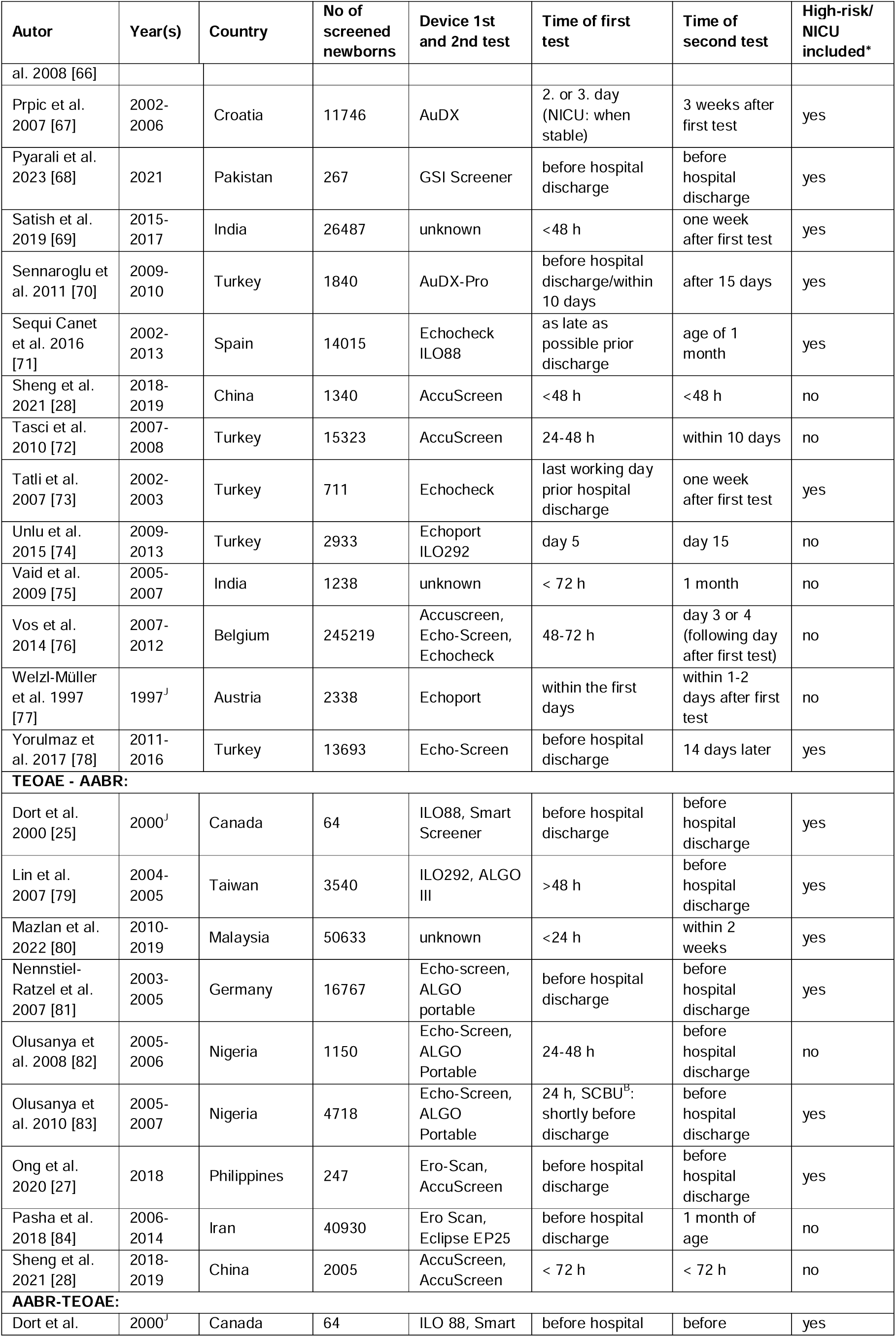
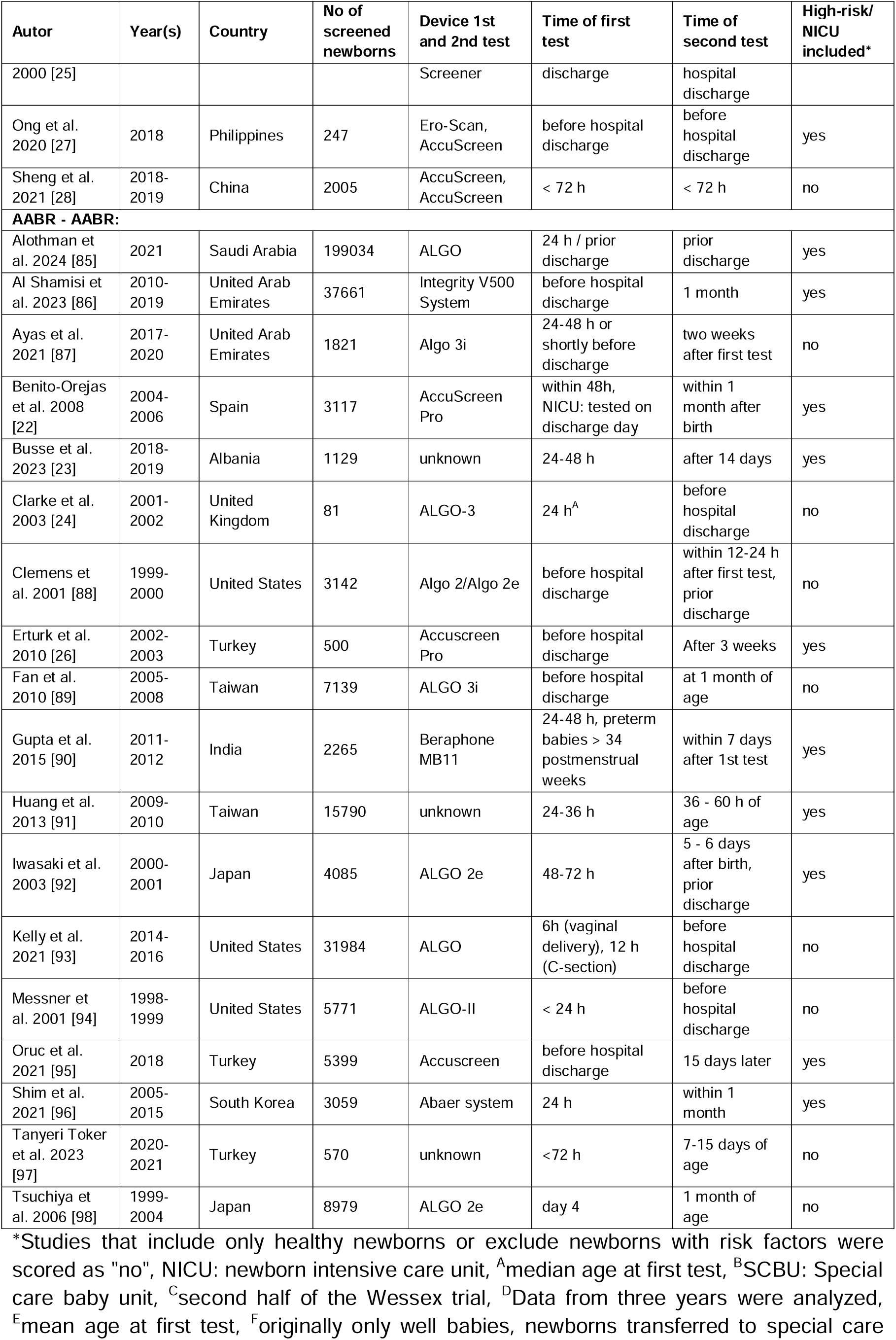
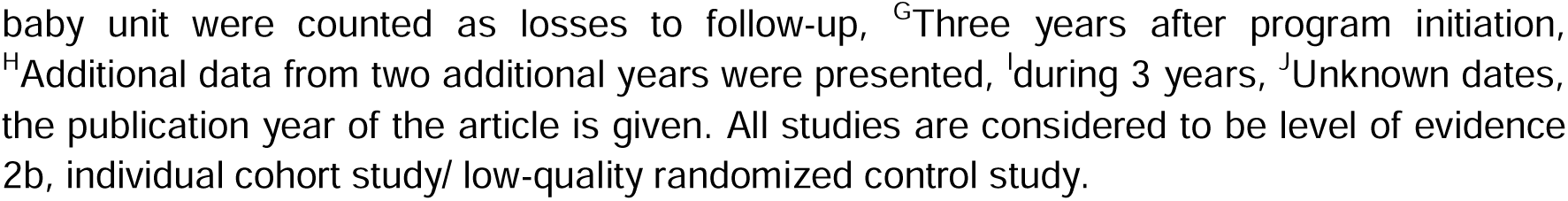
Characteristics of the included study protocols.

All included studies are cohort studies and have a level of evidence 2b, individual cohort study/ low-quality randomized control study, following definitions given in https://guides.library.stonybrook.edu/evidence-based-medicine/levels_of_evidence. Of the 85 study protocols, n = 55 (64.7%) studies examined the TEOAE-TEOAE test combination, n = 9 (10.6%) examined the TEOAE-AABR test combination n = 3 (3.5%) examined the AABR-TEOAE test combination, and n = 18 (21.2%) examined the AABR-AABR test combination. The median study size across all study protocols was n = 3238 newborns (min - max: 64 - 245219). The median study size for TEOAE-TEOAE was n = 3724 newborns (min - max: 81 - 245219), for TEOAE-AABR it was n = 3540 newborns (min - max: 64 - 50633), for AABR-TEOAE it was n = 247 newborns (min - max: 64 - 2005), and for AABR- AABR it was n = 3614 newborns (min - max: 81 - 199034).

Fig 2A presents the loss rates of newborns who did not pass the first test and did not attend the second test for all test combinations. Depending on the study, loss rates of up to 72% were reported. Median loss rates were 14% for TEOAE-TEOAE, 5% for TEOAE-AABR, and zero for AABR-TEOAE and AABR-AABR. Fig 2B shows the RFR of all study protocols. Median RFRs of 2.5%, 4.8%, 6.5% and 1.1% were found for the TEOAE-TEOAE, TEOAE- AABR, AABR-TEOAE, and AABR-AABR combinations, respectively. All but three studies [24,51,78], reported RFR below 10%. Out of the 85 study protocols, 57 (67.1%) showed a RFR below 4%.

**Fig 2.**
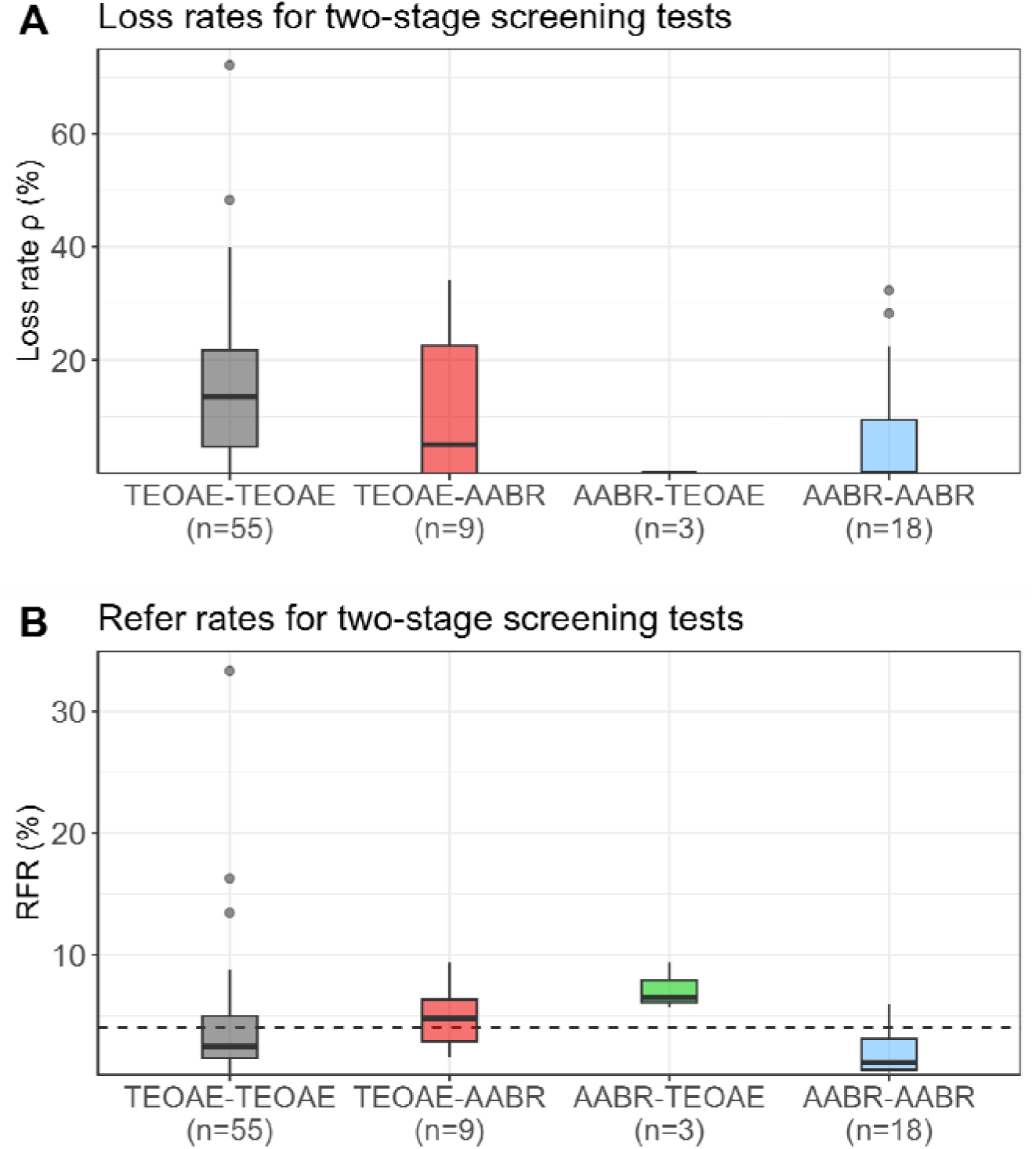
Loss rates (ρ) (A) and refer rates (RFR) (B) for the four different screening test combinations. The dashed horizontal line indicates the 4% threshold quality criteria defined in the Pediatrics Directive for the RFR. AABR = automated auditory brainstem response, TEOAE = transitory evoked otoacoustic emission.

### Random-effects meta-analysis of RFR

Forest plots of the random-effects meta-analysis of RFR are presented in Fig 3 for the TEOAE-TEOAE test combination and in Fig 4 for the other test combinations. A summary of the meta-analysis results is presented in Table 2. Forest plots for the meta-analysis of well-baby protocols are provided in the Supporting Information, S2 Fig and S3 Fig.

**Fig 3.**
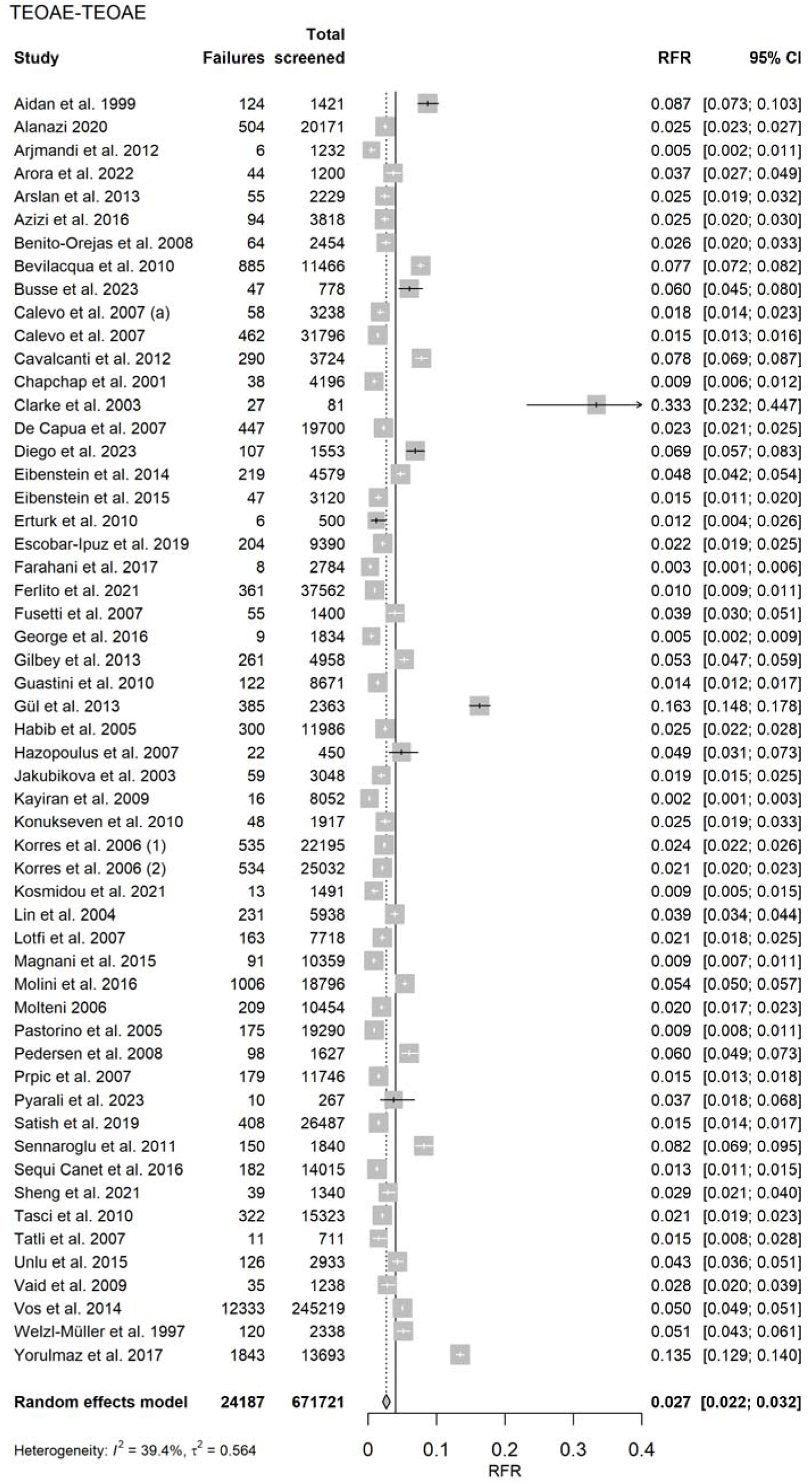
Random-effects meta-analysis of refer rate (RFR) for the 55 TEOAE-TEOAE study protocols. The table shows the number of newborns who did not pass the first and second test (“Failures”), the total number of screened newborns (“Total screened”), and the RFR with 95% confidence interval (95% CI) for each study. The summary estimate, including the 95% CI is shown as a gray diamond. The vertical solid line indicates the 4% threshold quality criteria defined in the Pediatrics Directive for the RFR. TEOAE = transitory evoked otoacoustic emission.

**Fig 4.**
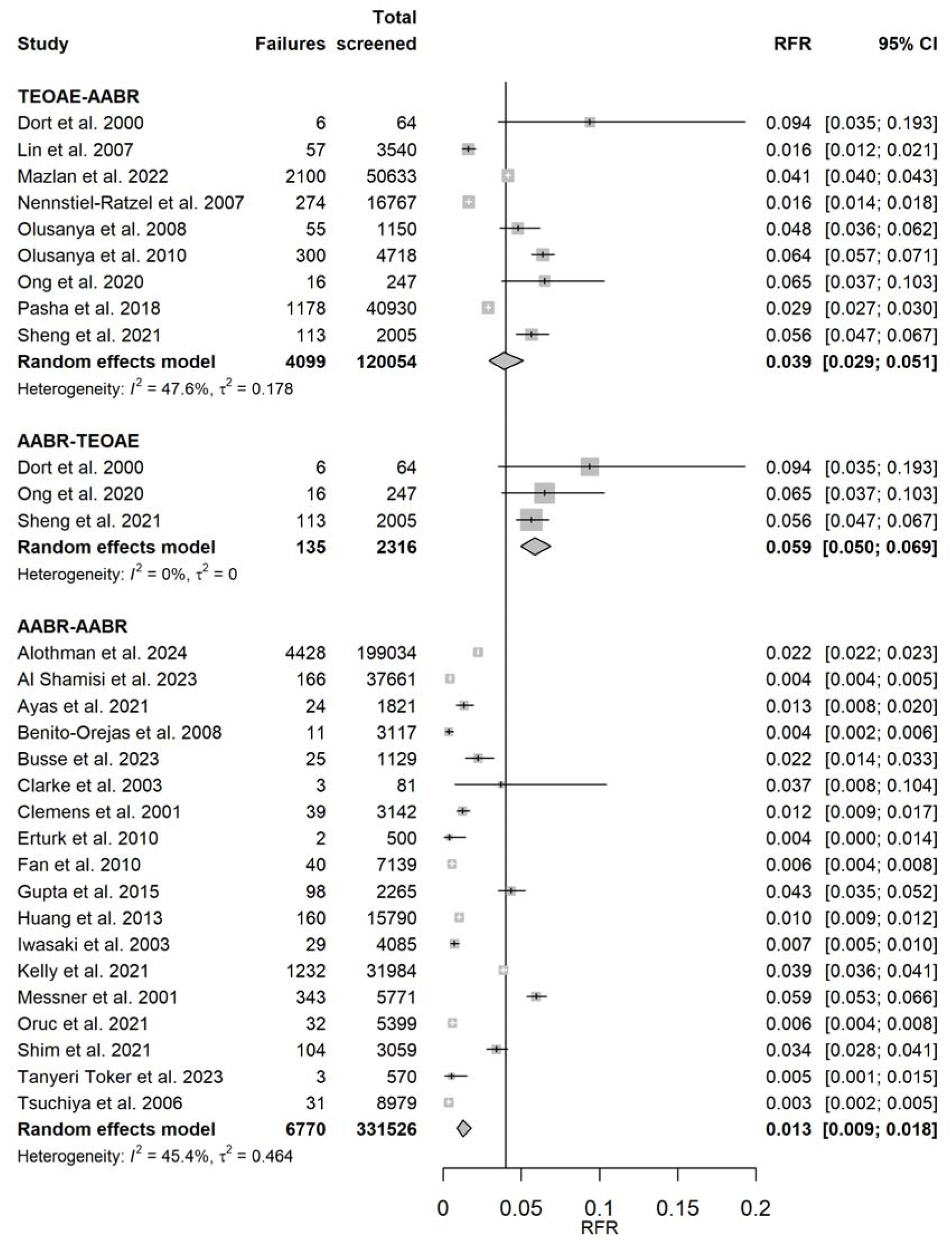
Random-effects meta-analysis of refer rate (RFR) for TEOAE-AABR, AABR- TEOAE, and AABR-AABR study protocols. The table shows the number of newborns who did not pass the first and second test (“Failures”), the total number of screened newborns (“Total screened”), and the RFR with 95% confidence interval (95% CI) for each study. The summary estimate per test combination, including the 95% CI, is shown as a gray diamond. The vertical solid line indicates the 4% threshold quality criteria defined in the Pediatrics Directive for the RFR. AABR = automated auditory brainstem response, TEOAE = transitory evoked otoacoustic emission.

**Table 2:** Results of the random effects meta-analysis of the refer rate.

| Test combination | RFR (95% CI) | I<sup>2</sup> | tau<sup>2</sup> | Q | df | p |
| --- | --- | --- | --- | --- | --- | --- |
| TEOAE-TEOAE | 2.7% (2.2, 3.2%) | 39.4% | 0.564 | 89.14 | 54 | **0.002** |
| TEOAE-AABR | 3.9% (2.9, 5.1%) | 47.6% | 0.178 | 15.27 | 8 | 0.054 |
| AABR-TEOAE | 5.9% (5.0, 6.9%) | 0% | 0 | 1.76 | 2 | 0.416 |
| AABR-AABR | 1.3% (0.9, 1.8%) | 45.4% | 0.464 | 31.14 | 17 | **0.019** |

RFR = refer rate, CI = confidence interval, AABR = automated auditory brainstem response, TEOAE = transitory evoked otoacoustic emission. The p-value shown in the right column relates to the Cochrane's Q statistic (Q) for heterogeneity. Statistically significant p values ( $p < 0.05$ ) are printed in bold.

Strategies that do not involve a change in the screening test method showed the lowest RFR. The upper limits of their 95% confidence intervals are below the recommended quality threshold of 4%. When the screening test method is changed between stage 1 and stage 2, the corresponding 95% confidence intervals cover or exceed the 4% threshold.

Studies of both screening combinations TEOAE-TEOAE and AABR-AABR show a moderate degree of heterogeneity as quantified by I^2 (39.4% and 45.4%, respectively). Excluding the TEOAE-TEOAE study protocol with a remarkably high RFR of 33.3% [24] and the two other studies with RFR > 10% [51,78], as shown in S4 Fig, reduces the summary estimate for RFR from 2.7% (2.2, 3.2%) to 2.4% (2.0, 2.8%). The results for RFR for well babies (S2 Fig and S3 Fig) are comparable to those of all studies: AABR-AABR and TEOAE-TEOAE show the lowest RFR (less than 4%). Changing the test method results in a higher RFR.

The results of the meta-regression of the failure rate of the first test and the RFR are presented in S1 Table and S5 Fig. The meta-regression regresses the RFR on the positive rate of the first stage (Eq.2) and allows inference on the conditional positivity rate in stage 2.

### Bias assessment

Fig 5 shows the results of the bias assessment. While there was a low risk of bias in the use of the tests (first test: 98.8%, second test: 100%), about half of the studies had a high risk of bias in patient selection and in flow and timing (49.4% and 52.9%, respectively). In terms of applicability concerns, about 3 out of 4 studies had high concerns for patient selection (72.9%). Similar to the risk of bias, both tests showed only low applicability concerns (first test: 97.6%, second test: 100%). The study-level bias assessment for all domains can be found in S2 Table.

**Fig 5.**
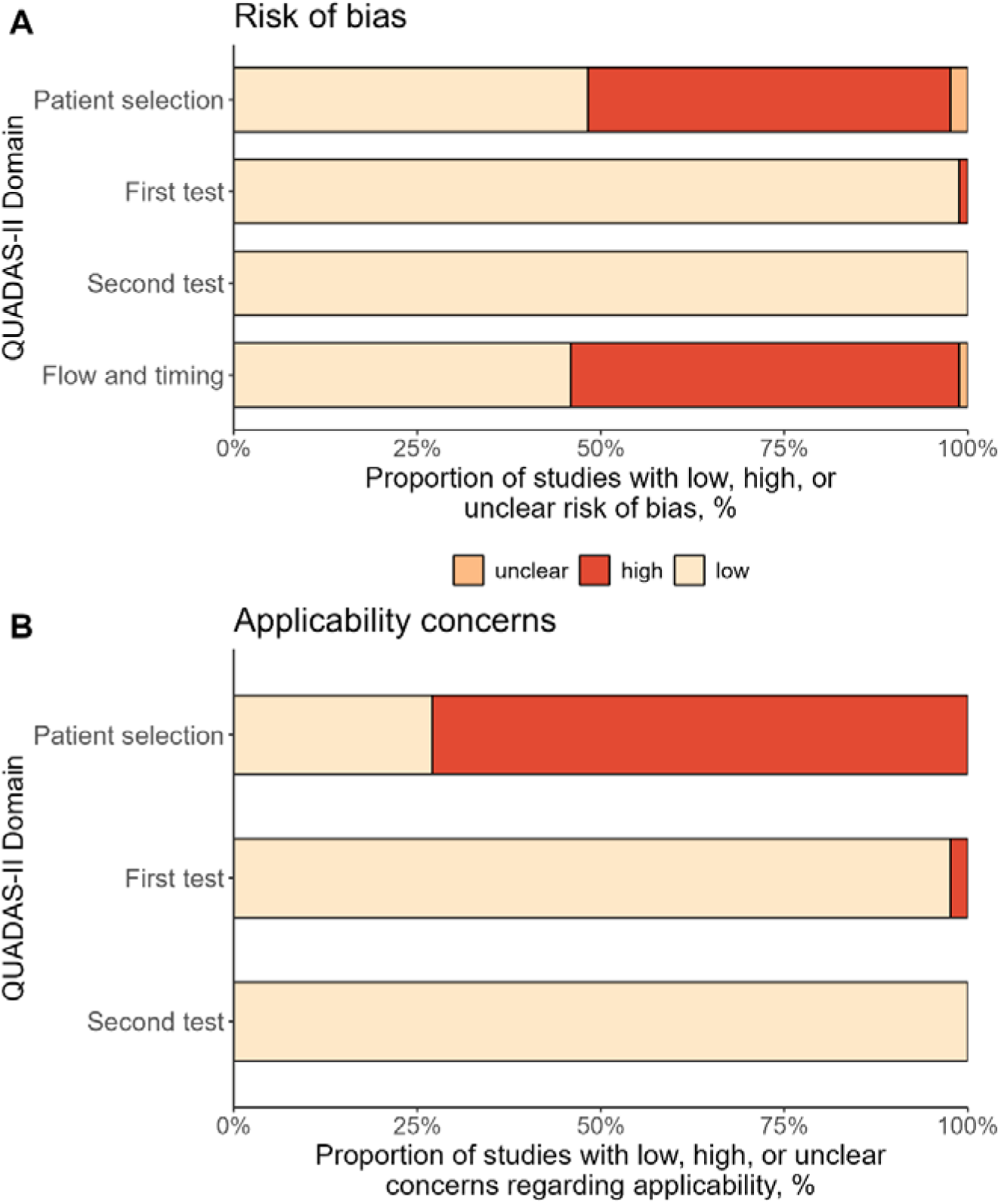
Bias assessment using QUADAS-II tool. The figure shows the percentage of the included 85 study protocols with low, high, or unclear risk of bias (A) and concerns regarding applicability (B).

The GRADE Summary of Findings table is included in the S1 File. We rated all evidence generated in this meta-analysis as moderate, which is the second highest GRADE evidence category. The level of evidence for the RFR from the random-effects meta-analysis was lowered to moderate because of the moderate heterogeneity found, and for test combinations with test method change between the two stages because of the small number of pooled studies.

## Discussion

A meta-analysis was conducted to identify the optimal screening algorithm for a two-stage NHS using combinations of TEOAE or AABR tests. The study analyzed 85 study protocols with over 1,12 million newborns meeting the inclusion criteria. The results showed that the refer rate (RFR) was lower when there was no change in the screening method used. The aggregated RFR was 1.3% for the AABR-AABR test combination and 2.7% for TEOAE- TEOAE.

The following discussion focuses on newborn hearing screening in the German setting. However, we believe that the discussion will be of broader interest to other countries following similar screening strategies, as the hearing screening process should always be evaluated and adjusted as necessary to achieve the best possible quality.

The RFR is an important quality parameter in the NHS, as screenings with a refer result must be followed up by a pediatric audiologist. These specialists are scarce in Germany (as well as in other countries), which can lead to long waiting times for families with children who have not passed the screening tests. In addition, false-positive results can cause unnecessary anxiety for parents [99]. A low RFR in the NHS can be achieved primarily through a multi-step screening algorithm. Therefore, when the NHS was introduced in 2009, the German Pediatrics Directive required an AABR control if the first hearing test was not passed and an RFR of less than 4% at discharge, in line with national and international quality targets at the time [14,100]. The UK even requested lower RFRs (acceptable: 3%, achievable: 2%) [17].

The German NHS evaluation data for the years 2011/2012 and the follow-up evaluation data for 2017/2018 both indicate that a second hearing test following an initial screening test with a “fail” result considerably reduces the RFR, as this second test was passed in over 80% [8,15]. However, it is worth noting, that this second measurement was performed with a TEOAE in more than half of the tests, which is contrary to the German Pediatrics Directive. In follow-up evaluation interviews this was explained by the longer measurement duration and the increased susceptibility to interference of the AABR. The follow-up evaluation analysis revealed a considerably higher rate of refer results in the second test when a different test method was used than in the first test [8,15]. This is in line with the finding of this meta-analysis that the highest RFR were observed when the test method was changed (TEOAE-AABR or AABR-TEOAE).

In the German follow-up evaluation, the TEOAE-TEOAE algorithm had a lower RFR (9.62%) than AABR-AABR (13.98%) [15]. In contrast, in this meta-analysis the AABR-AABR test combination had the lowest RFR, even lower than that of the TEOAE-TEOAE algorithm. This was also observed in the subgroup analysis on “well babies” and may be attributed to the higher lost-to-follow-up rate in the studies reporting TEOAE-TEOAE results (see Fig 2). In addition, studies utilizing this test combination exhibited a significantly higher heterogeneity (see Table 2, Q statistic) and were often based on routine clinical data, whereas data for the AABR-AABR test sequence were mainly derived from clinical studies. The AABR diagnostic is the ”gold standard” for detecting most hearing disorders. However, since TEOAE offers the most practical screening setting and TEOAE-TEOAE has the second best refer rate, the authors recommend this combination for well-babies. TEOAE is known to miss retrocochlear causes of hearing loss, such as auditory neuropathy. However, this condition is rare in well babies. Retrocochlear hearing loss is most common in children with risk factors for hearing disorders such as hyperbilirubinemia.

One limitation of our study is the assumption of homogeneity of sensitivities and specificities across all studies, which ignores the heterogeneity caused by differently qualified personnel and in different settings (i.e. quiet vs. noisy). As these factors influencing the screening result are only described in detail in a few reports, they could not be considered in the meta-analysis. Similarly, the reports often lack information on whether data from children with risk factors for hearing impairment were included. However, they usually provide information on whether children from the NICU were included. In the subgroup analysis including exclusively study protocols with newborns without risk factors (“well babies”), the AABR- AABR and TEOAE-TEOAE algorithms showed the lowest RFR (below 4%) as well as higher RFR after changing the test method.

To our knowledge, this is the first systematic review on quality measures of tests used in a two-stage NHS. Strengths of the study include the large number of newborns included and the high number of study reports. While we analyzed the refer rate using a standard frequentist random-effects meta-analysis, our proposed method of estimating the RFR using Bayesian methods may provide an interesting way for analysis in future studies.

In its most recent 2019 position paper, the Joint Committee on Infant Hearing recommends performing at least two screening attempts with the same method or an AABR after TEOAE before discharge of a well baby. TEOAE testing after an initial AABR with a refer result is also acceptable for well babies, as the lost-to-follow-up rate for outpatient follow-up is very high [6,101]. The UK screening program guidelines recommend performing a further TEOAE test with an interval of at least 5 hours if the first TEOAE test is not passed for well babies [17].

The results of the meta-analysis and the data analysis of the German follow-up evaluation should provide evidence for adjusting the German Pediatrics Directive regarding the method of the second test to improve the RFR and align with international recommendations. Staff compliance with performing a second test before discharge is expected to improve if TEOAE tests are allowed, as TEOAE tests are faster and easier to perform than an AABR measurement. In contrast, changing the method of the second hearing test after failing the initial test results in a higher RFR without evident advantages. However, in children with risk factors for perinatal hearing impairment, both hearing tests should always be performed with an AABR test, as specified in the German Pediatrics Directive [14] and international guidelines.

## Data Availability

All data produced are available online at https://osf.io/nuk4p/.

## Acknowledgments

We would like to thank Anja Friedrichs for the full-text retrieval of non-open access studies.

## Supporting Information

**S1 Fig.**
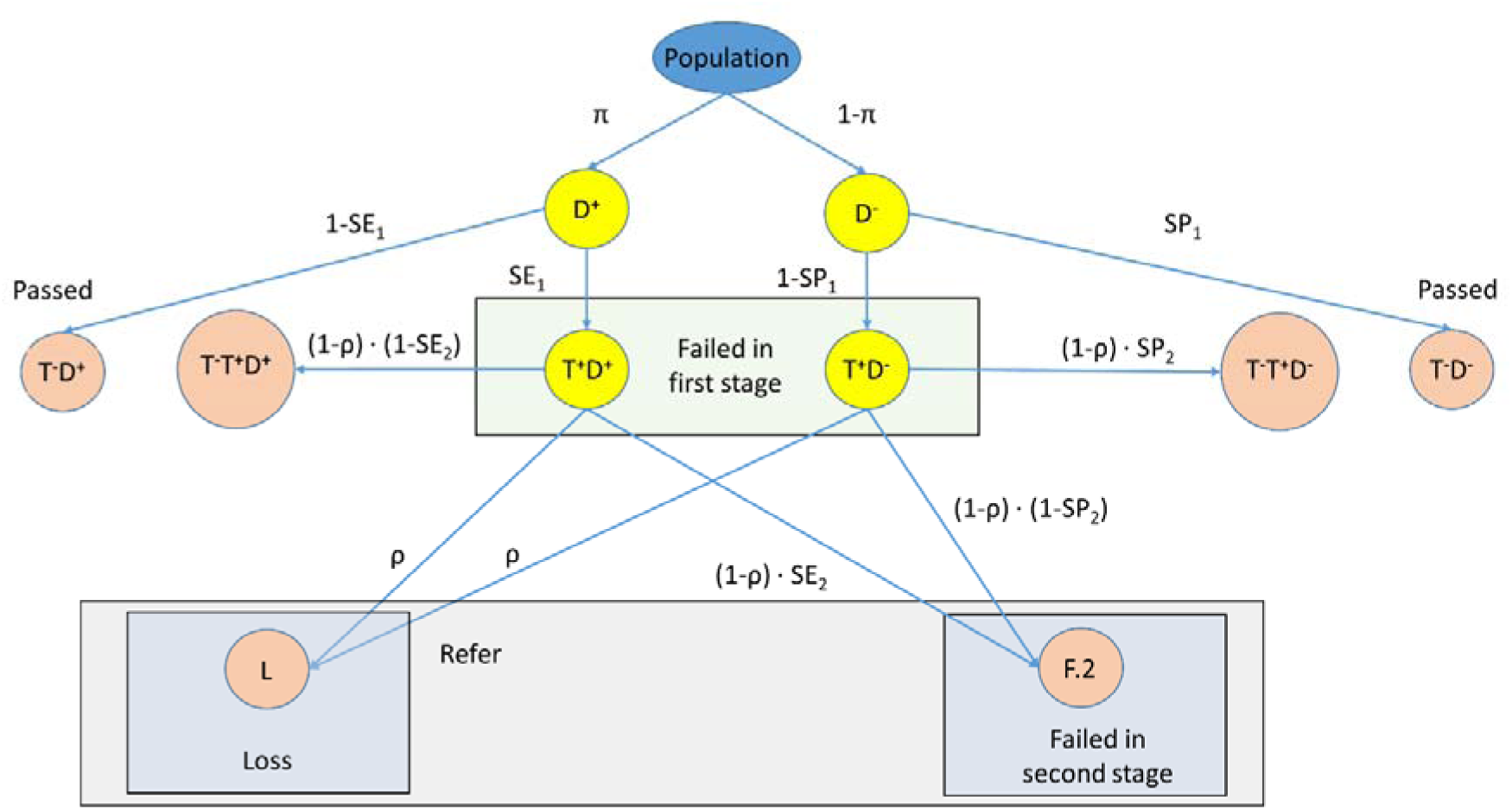
Model for two-stage newborn hearing screening. π = prevalence of hearing impairment, D+ (D-) = newborns with (without) hearing impairment, T- (T+) =newborns with “pass” (“fail”) test results, SE_1_ (SE_2_) = sensitivity of the first (second) test, SP_1_ (SP_2_) = specificity of the first (second) test, ρ = loss rate after the first test stage, L = number of newborns lost after the first test result was “fail”, F.2=number of newborns whose second test result was “fail”.

### S1 Checklist. PRISMA checklist

**S1 Checklist. PRISMA checklist.**
| Section and Topic | Item # | Checklist item | Location where item is reported |
| --- | --- | --- | --- |
| **TITLE** |  |  |  |
| Title | 1 | Identify the report as a systematic review. | Title (p. 1) |
| **ABSTRACT** |  |  |  |
| Abstract | 2 | See the PRISMA 2020 for Abstracts checklist. | Considered in abstract (p. 2) |
| **INTRODUCTION** |  |  |  |
| Rationale | 3 | Describe the rationale for the review in the context of existing knowledge. | Introduction paragraphs 1-4 (pp.3-4 ) |
| Objectives | 4 | Provide an explicit statement of the objective(s) or question(s) the review addresses. | Introduction paragraph 4 (p. 4) |
| **METHODS** |  |  |  |
| Eligibility criteria | 5 | Specify the inclusion and exclusion criteria for the review and how studies were grouped for the syntheses. | Materials and methods → Eligibility criteria (pp. 7-8) |
| Information sources | 6 | Specify all databases, registers, websites, organisations, reference lists and other sources searched or consulted to identify studies. Specify the date when each source was last searched or consulted. | Materials and methods → Search method (p. 8) |
| Search | 7 | Present the full search strategies for all databases, | Materials and methods |
| strategy |  | registers and websites, including any filters and limits used. | → Search method (p. 8) |
| Selection process | 8 | Specify the methods used to decide whether a study met the inclusion criteria of the review, including how many reviewers screened each record and each report retrieved, whether they worked independently, and if applicable, details of automation tools used in the process. | Materials and methods<br>→ Study records (pp. 8-9) |
| Data collection process | 9 | Specify the methods used to collect data from reports, including how many reviewers collected data from each report, whether they worked independently, any processes for obtaining or confirming data from study investigators, and if applicable, details of automation tools used in the process. | Materials and methods<br>→ Data extraction and items (pp. 9-10) |
| Data items | 10a | List and define all outcomes for which data were sought. Specify whether all results that were compatible with each outcome domain in each study were sought (e.g. for all measures, time points, analyses), and if not, the methods used to decide which results to collect. | Materials and methods<br>→ Data extraction and items (pp. 9-10) |
|  | 10b | List and define all other variables for which data were sought (e.g. participant and intervention characteristics, funding sources). Describe any assumptions made about any missing or unclear information. | Materials and methods<br>→ Data extraction and items (pp. 9-10), legend of Table 1 (p. 19) |
| Study risk of bias assessment | 11 | Specify the methods used to assess risk of bias in the included studies, including details of the tool(s) used, how many reviewers assessed each study and whether they worked independently, and if applicable, details of automation tools used in the process. | Materials and methods<br>→ Risk of bias in individual studies and publication bias (pp. 10-11) |
| Effect measures | 12 | Specify for each outcome the effect measure(s) (e.g. risk ratio, mean difference) used in the synthesis or presentation of results. | Materials and methods<br>→ Data extraction and items (pp. 9-10) |
| Synthesis methods | 13a | Describe the processes used to decide which studies were eligible for each synthesis (e.g. tabulating the study intervention characteristics and comparing against the planned groups for each synthesis (item #5)). | Materials and methods<br>→ Statistical analysis (p. 11) |
|  | 13b | Describe any methods required to prepare the data for presentation or synthesis, such as handling of missing summary statistics, or data conversions. | Materials and methods<br>→ Data extraction and items (pp. 9-10) |
|  | 13c | Describe any methods used to tabulate or visually display results of individual studies and syntheses. | Materials and methods<br>→ Statistical analysis (p. 11) |
|  | 13d | Describe any methods used to synthesize results and provide a rationale for the choice(s). If meta-analysis was performed, describe the model(s), method(s) to identify the presence and extent of statistical heterogeneity, and software package(s) used. | Materials and methods<br>→ Statistical analysis (p. 11) |
|  | 13e | Describe any methods used to explore possible causes of heterogeneity among study results (e.g. subgroup analysis, meta-regression). | Materials and methods<br>→ Subgroup analysis (p. 11) |
|  | 13f | Describe any sensitivity analyses conducted to assess robustness of the synthesized results. | Materials and methods<br>→ Subgroup analysis (p. 11) |
| Reporting bias assessment | 14 | Describe any methods used to assess risk of bias due to missing results in a synthesis (arising from reporting biases). | Materials and methods<br>→ Risk of bias in individual studies and publication bias (pp. 10-11) |
| Certainty assessment | 15 | Describe any methods used to assess certainty (or confidence) in the body of evidence for an outcome. | Materials and methods<br>→ Confidence in cumulative evidence (p. 11) |
| **RESULTS** |  |  |  |
| Study selection | 16a | Describe the results of the search and selection process, from the number of records identified in the search to the number of studies included in the review, ideally using a flow diagram. | Fig 1 (p. 12) |
|  | 16b | Cite studies that might appear to meet the inclusion criteria, but which were excluded, and explain why they were excluded. | Legend of Fig 1 (p. 11) |
| Study characteristics | 17 | Cite each included study and present its characteristics. | Table 1 (pp. 13-19) |
| Risk of bias in studies | 18 | Present assessments of risk of bias for each included study. | S2 Table (p.29 ) |
| Results of individual studies | 19 | For all outcomes, present, for each study: (a) summary statistics for each group (where appropriate) and (b) an effect estimate and its precision (e.g. confidence/credible interval), ideally using structured tables or plots. | Fig 2 (p. 20) |
| Results of syntheses | 20a | For each synthesis, briefly summarise the characteristics and risk of bias among contributing studies. | Fig 3 (p. 20) |
|  | 20b | Present results of all statistical syntheses conducted. If meta-analysis was done, present for each the summary estimate and its precision (e.g. confidence/credible interval) and measures of statistical heterogeneity. If comparing groups, describe the direction of the effect. | See Figs 3 and 4 (p. 20, p. 21) and Tables 2 and 3 (p. 21, p. 23) |
|  | 20c | Present results of all investigations of possible causes of heterogeneity among study results. | S4 Fig (pp. 28-29) |
|  | 20d | Present results of all sensitivity analyses conducted to assess the robustness of the synthesized results. | S4 Fig (pp. 28-29) |
| Reporting biases | 21 | Present assessments of risk of bias due to missing results (arising from reporting biases) for each synthesis assessed. | NA, since no reporting bias occurred. |
| Certainty of evidence | 22 | Present assessments of certainty (or confidence) in the body of evidence for each outcome assessed. | S1 File (p. 29) |
| **DISCUSSION** |  |  |  |
| Discussion | 23a | Provide a general interpretation of the results in the context of other evidence. | Discussion paragraphs 3-5 (pp. 25-26) |
|  | 23b | Discuss any limitations of the evidence included in the review. | Discussion paragraph 6 (p. 26) |
|  | 23c | Discuss any limitations of the review processes used. | Discussion paragraph 7 (p. 26) |
|  | 23d | Discuss implications of the results for practice, policy, and future research. | Discussion last paragraph (p. 27) |
| **OTHER INFORMATION** |  |  |  |
| Registration and protocol | 24a | Provide registration information for the review, including register name and registration number, or state that the review was not registered. | Materials and methods<br>→ Study protocol (p. 7) |
|  | 24b | Indicate where the review protocol can be accessed, or state that a protocol was not prepared. | Materials and methods<br>→ Study protocol (p. 7) |
|  | 24c | Describe and explain any amendments to information provided at registration or in the protocol. | Materials and methods<br>→ Study protocol (p. 7) |
| Support | 25 | Describe sources of financial or non-financial support for the review, and the role of the funders or sponsors in the review. | See Additional information |
| Competing interests | 26 | Declare any competing interests of review authors. | See Additional information |
| Availability of data, code and other materials | 27 | Report which of the following are publicly available and where they can be found: template data collection forms; data extracted from included studies; data used for all analyses; analytic code; any other materials used in the review. | Materials and Methods<br>→ Statistical analysis (p. 11) |

NA = not applicable.

**S2 Fig.**
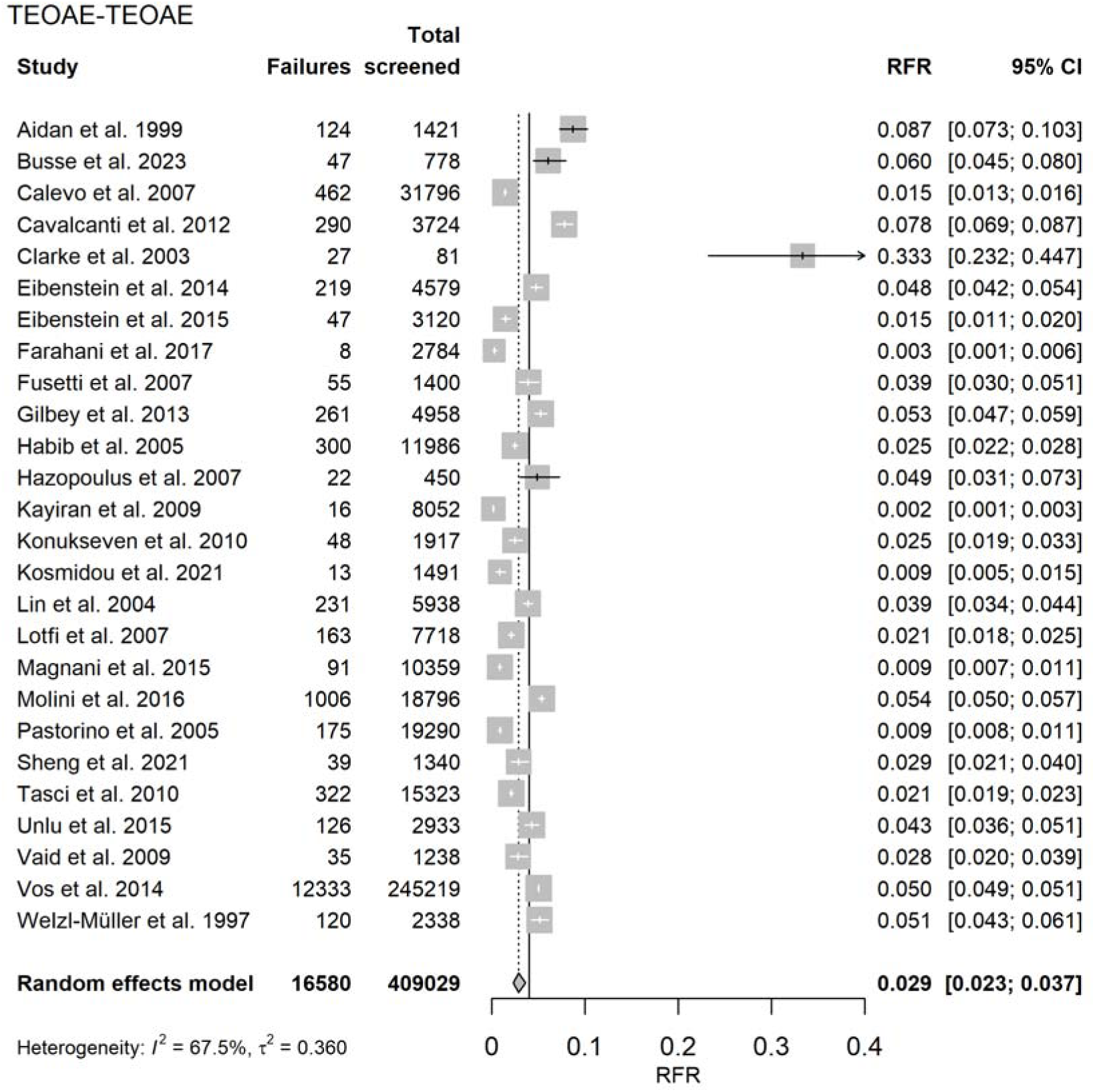
Random-effects meta-analysis of refer rate (RFR) for the 26 TEOAE-TEOAE well-baby study protocols. Shown are the number of newborns who failed both the first and second test (“Failures”), the number of screened newborns (“Total screened”), and the RFR with the 95% confidence interval (95% CI) for each study. The summary estimate including the 95% CI is shown as a gray diamond. The vertical solid line indicates the 4% threshold quality criteria defined in the Pediatrics Directive for the RFR. TEOAE = transitory evoked otoacoustic emission.

**S3 Fig.**
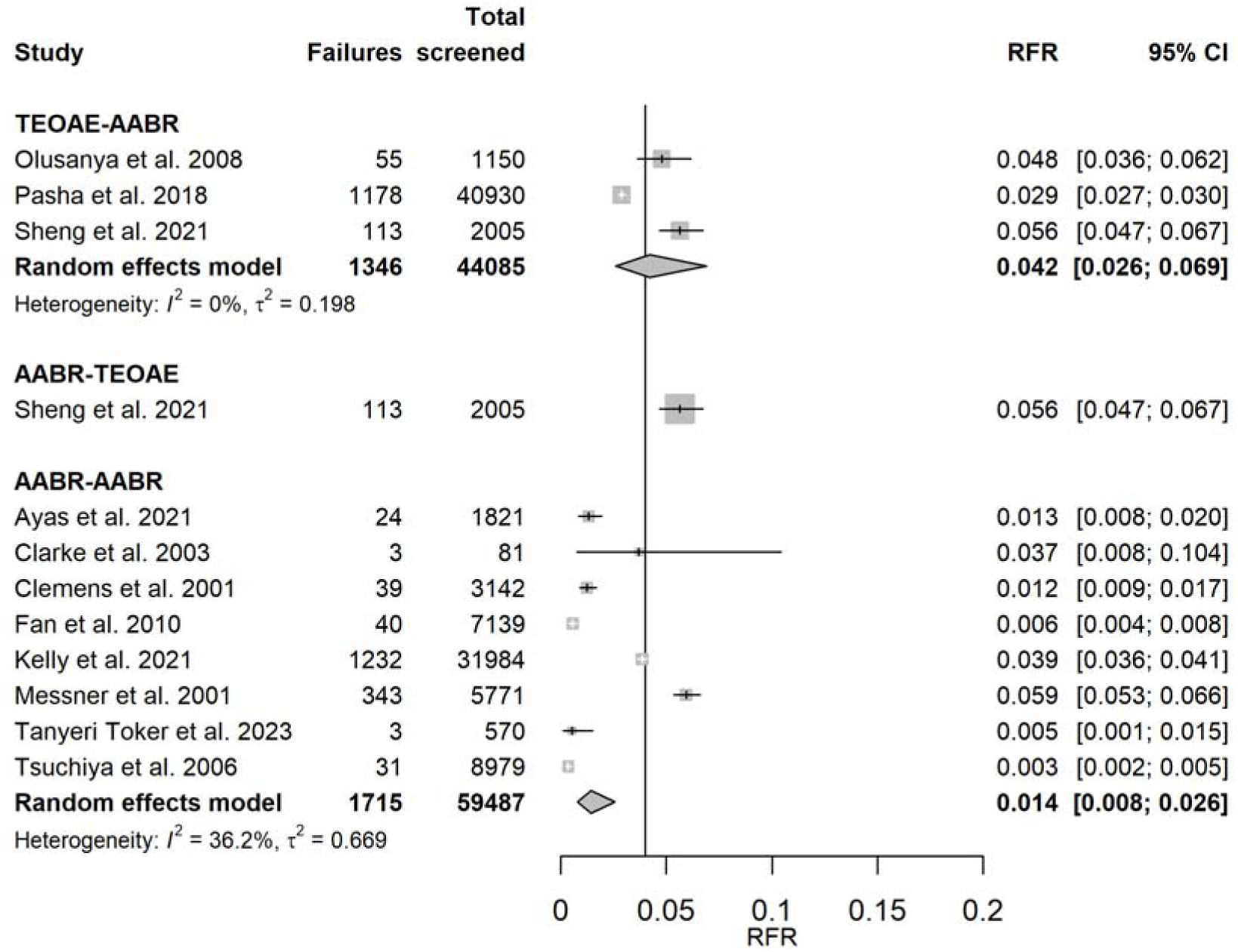
Random-effects meta-analysis of refer rate (RFR) for TEOAE-AABR, AABR- TEOAE and AABR-AABR well-baby study protocols. Shown are the number of newborns who did not pass first and second test (“Failures”), the number of screened newborns (“Total screened”), and the RFR with the 95% confidence interval (95% CI) for each study. The summary estimate per test combination including the 95% CI is shown as a gray diamond. The vertical solid line indicates the 4% threshold quality criteria defined in the Pediatrics Directive for the RFR. AABR = automated auditory brainstem response, TEOAE = transitory evoked otoacoustic emission.

**S4 Fig.**
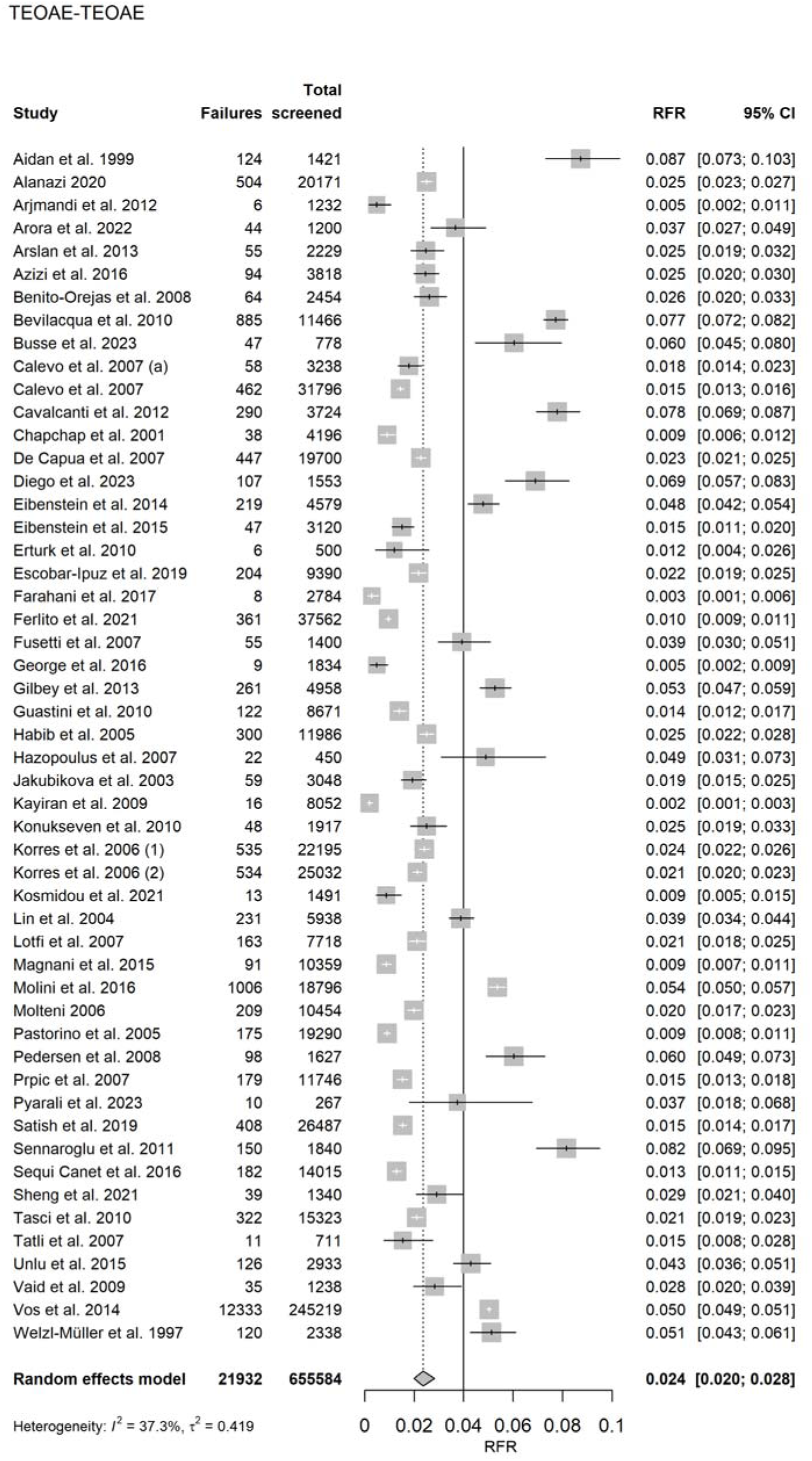
Random-effects meta-analysis of refer rate (RFR) for 52 TEOAE-TEOAE study protocols without outliers. Excluding the studies by Clarke et al. 2003, Gül et al. 2013 and Yorulmaz et al. 2017. Shown are the number of newborns who did not pass first and second test (“Failures”), the number of screened newborns (“Total screened”), and the RFR with 95% confidence interval (95% CI) for each study. The summary estimate including the 95% CI is shown as a gray diamond. The vertical solid line indicates the 4% threshold quality criteria defined in the Pediatrics Directive for the RFR. TEOAE = transitory evoked otoacoustic emission.

**Table S1:**
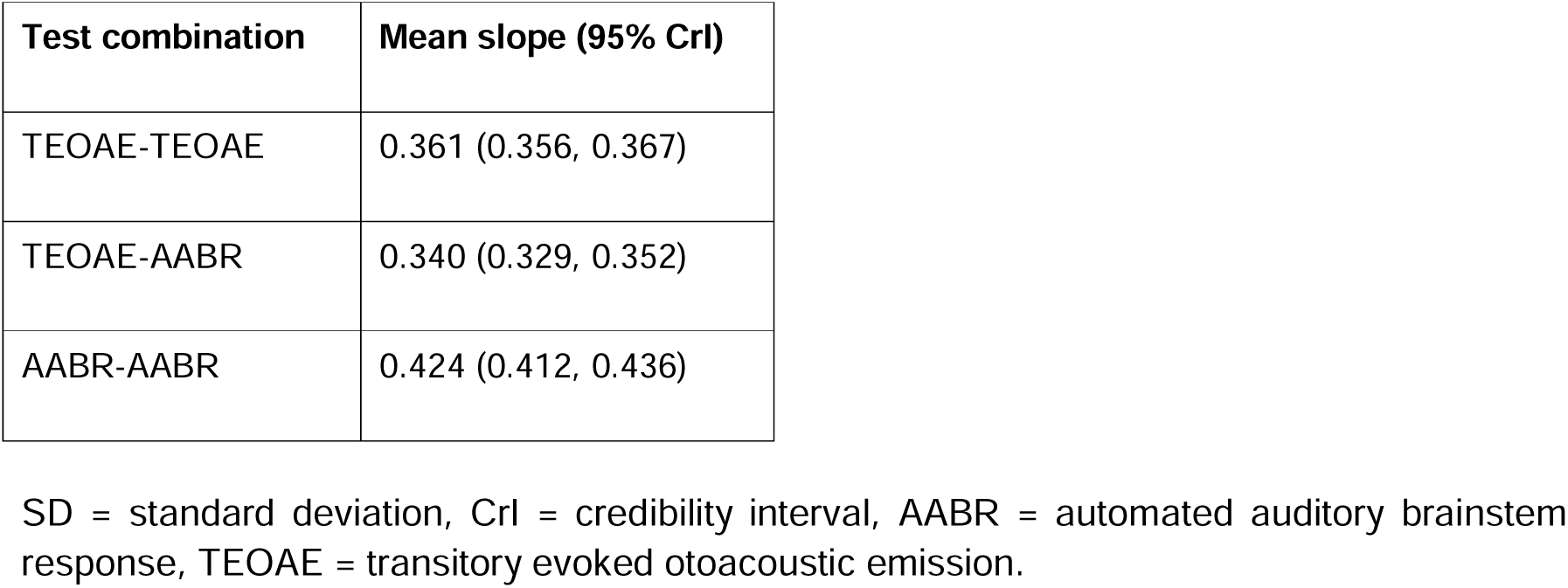
Results of the meta-regression.

| Test combination | Mean slope (95% CrI) |
| --- | --- |
| TEOAE-TEOAE | 0.361 (0.356, 0.367) |
| TEOAE-AABR | 0.340 (0.329, 0.352) |
| AABR-AABR | 0.424 (0.412, 0.436) |

SD = standard deviation, CrI = credibility interval, AABR = automated auditory brainstem response, TEOAE = transitory evoked otoacoustic emission.

**S5 Fig.**
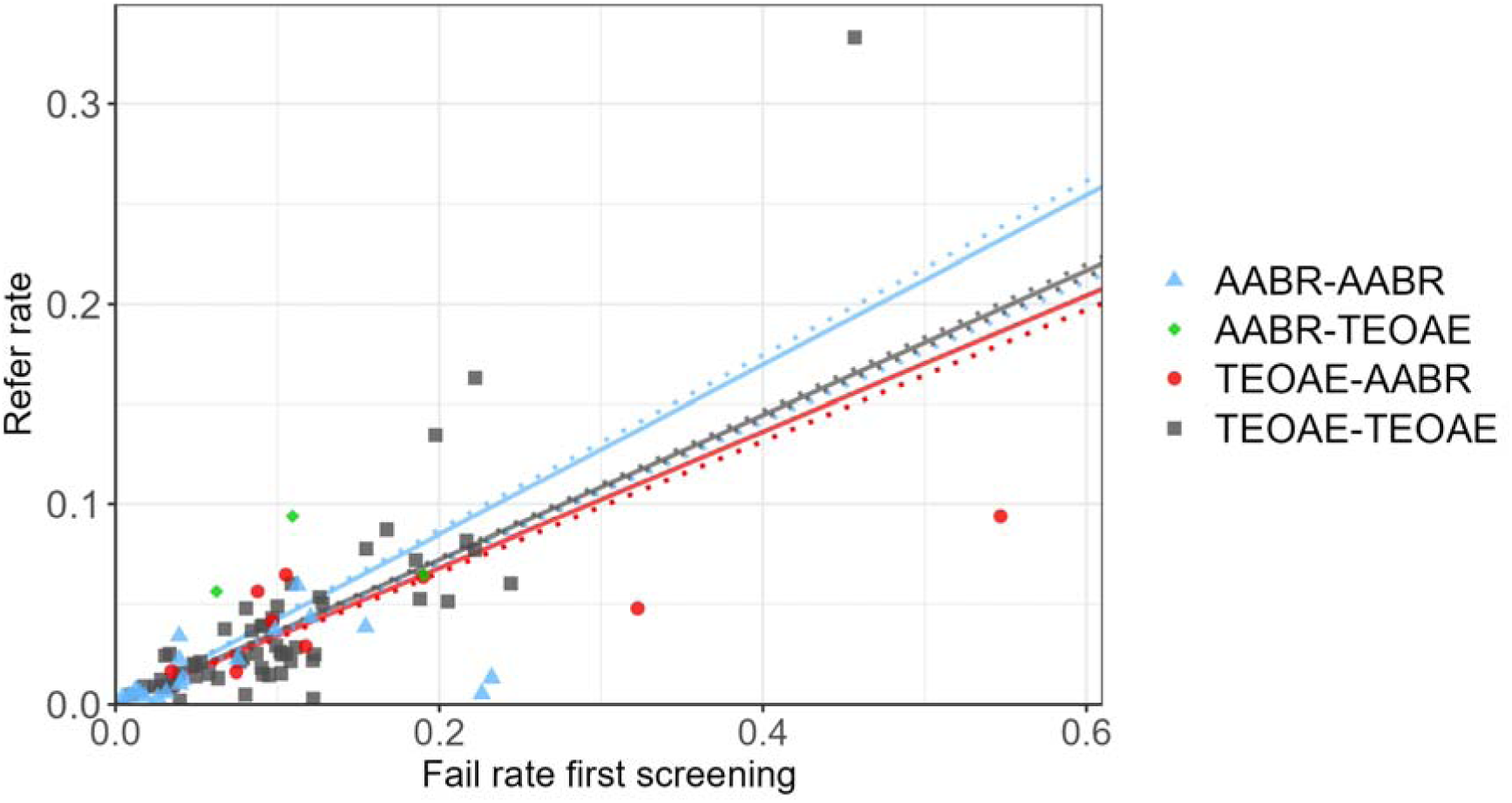
Refer rates in dependence of the failure rate of the first test for the test combinations AABR-AABR (n=18, blue triangles), AABR-TEOAE (n=3, green diamonds), TEOAE-AABR (n=9, red dots) and TEOAE-TEOAE (n=55, gray squares). AABR = automated auditory brainstem response, TEOAE = transitory evoked otoacoustic emission. Meta-regression lines including credibility intervals are plotted with data from S1 Table. No meta-regression line is shown for the AABR-TEOAE test combination with only three studies

**Table S2:** Study-level assessment of bias for all 85 study protocols.

|  | Risk of bias |  |  |  | Applicability concerns |  |  |
| --- | --- | --- | --- | --- | --- | --- | --- |
| Study | Patient selection | First test | Second test | Flow and timing | Patient selection | First test | Second test |
| **TEOAE-TEOAE:** |  |  |  |  |  |  |  |
| Aidan et al. 1999 | low | low | low | high | high | low | low |
| Alanazi 2020 | low | low | low | high | high | low | low |
| Arjmandi et al. 2012 | high | low | low | high | high | Low | low |
| Arora et al. 2022 | high | low | low | high | high | low | low |
| Arslan et al. 2013 | high | low | low | low | high | low | low |
| Azizi et al. 2016 | high | low | low | high | high | low | low |
| Benito-Orejas et al. 2008 | high | low | low | high | high | low | low |
| Bevilacqua et al. 2021 | low | low | low | high | high | low | low |
| Busse et al. 2023 | low | low | low | high | high | low | low |
| Calevo et al. 2007 (a) | unclear | low | low | low | low | low | low |
| Calevo et al. 2007 | low | low | low | low | low | low | low |
| Calvacanti et al. 2012 | high | low | low | high | high | low | low |
| Chapchap et al. 2001 | high | low | low | high | high | low | low |
| Clarke et al. 2003 | high | low | low | low | low | low | low |
| De Capua et al. 2007 | high | low | low | high | high | low | low |
| Diego et al. 2023 | high | low | low | high | high | low | low |
| Eibenstein et al. 2014 | low | low | low | high | high | low | low |
| Eibenstein et al 2015 | low | low | low | high | low | low | low |
| Erturk et al. 2010 | unclear | low | low | high | high | low | low |
| Escobar-Ipuz et al. 2019 | high | low | low | low | high | low | low |
| Farahani et al. 2017 | low | low | low | low | low | low | low |
| Ferlito et al. 2021 | high | low | low | high | high | low | low |
| George et al. 2016 | high | low | low | high | high | low | low |
| Fusetti et al. 2007 | low | low | low | high | high | low | low |
| Gilbey et al. 2013 | low | low | low | low | low | low | low |
| Guastini et al. 2010 | high | low | low | low | high | low | low |
| Study | Patient selection | First test | Second test | Flow and timing | Patient selection | First test | Second test |
| Gül et al. 2013 | high | low | low | high | high | low | low |
| Habib et al. 2005 | low | low | low | low | low | low | low |
| Hatzopoulus et al. 2007 | low | low | low | high | high | low | low |
| Jakubikova et al. 2003 | high | low | low | high | high | low | low |
| Kayiran et al. 2009 | high | low | low | low | high | low | low |
| Konukseven et al. 2010 | low | low | low | high | high | low | low |
| Korres et al. 2006 (1) | high | high | low | high | high | high | low |
| Korres et al. 2006 (2) | low | low | low | low | low | low | low |
| Kosmidou et al. 2021 | low | low | low | low | low | low | low |
| Lin et al. 2004 | high | low | low | high | high | low | low |
| Lotfi et al. 2007 | low | low | low | high | low | low | low |
| Magnani et al. 2015 | low | low | low | low | low | low | low |
| Molini et al. 2016 | low | low | low | high | high | low | low |
| Molteni 2006 | high | low | low | high | high | low | low |
| Pastorino et al. 2005 | low | low | low | high | high | high | low |
| Pedersen et al. 2008 | high | low | low | high | high | low | low |
| Prpic et al. 2007 | high | low | low | low | high | low | low |
| Pyrali et al. 2023 | high | low | low | low | high | low | low |
| Satish et al. 2019 | high | low | low | low | high | low | low |
| Sennaroglu et al. 2011 | high | low | low | unclear | high | low | low |
| Sequi Canet et al. 2016 | low | low | low | high | high | low | low |
| Sheng et al. 2021 | low | low | low | high | high | low | low |
| Tasci et al. 2010 | low | low | low | low | high | low | low |
| Tatli et al. 2007 | high | low | low | high | high | low | low |
| Unlu et al. 2015 | low | low | low | low | low | low | low |
| Vaid et al. 2009 | low | low | low | high | high | low | low |
| Vos et al. 2014 | low | low | low | high | high | low | low |
| Welzl-Müller et al. 1997 | low | low | low | high | high | low | low |
| Yorulmaz et al. 2017 | low | low | low | high | high | low | low |
| Study | Patient selection | First test | Second test | Flow and timing | Patient selection | First test | Second test |
| **TEOAE-AABR:** |  |  |  |  |  |  |  |
| Dort et al. 2000 | high | low | low | low | high | low | low |
| Lin et al. 2007 | low | low | low | low | low | low | low |
| Mazlan et al. 2022 | low | low | low | high | high | low | low |
| Nennstiel-Ratzel et al. 2007 | high | low | low | high | high | low | low |
| Olusanya et al. 2008 | low | low | low | low | low | low | low |
| Olusanya et al. 2010 | high | low | low | high | high | low | low |
| Ong et al. 2020 | high | low | low | low | high | low | low |
| Pasha et al. 2018 | low | low | low | high | high | low | low |
| Sheng et al. 2021 | low | low | low | low | low | low | low |
| **AABR-TEOAE:** |  |  |  |  |  |  |  |
| Dort et al. 2000 | high | low | low | low | high | low | low |
| Ong et al. 2020 | low | low | low | low | high | low | low |
| Sheng et al. 2021 | low | low | low | low | low | low | low |
| **AABR-AABR:** |  |  |  |  |  |  |  |
| Alothman et al. 2024 | high | low | low | low | high | low | low |
| Al Shamisi et al. 2023 | high | low | low | high | high | low | low |
| Ayas et al. 2021 | low | low | low | low | low | low | low |
| Benito-Orejas et al. 2008 | low | low | low | high | high | low | low |
| Busse et al. 2023 | high | low | low | low | high | low | low |
| Clarke et al. 2003 | high | low | low | low | low | low | low |
| Clemens et al. 2001 | low | low | low | low | low | low | low |
| Erturk et al. 2010 | high | low | low | high | high | low | low |
| Fan et al. 2010 | high | low | low | low | high | low | low |
| Gupta et al. 2015 | high | low | low | low | high | low | low |
| Huang et al. 2013 | high | low | low | low | high | low | low |
| Iwasaki et al. 2003 | low | low | low | low | low | low | low |
| Study | Patient selection | First test | Second test | Flow and timing | Patient selection | First test | Second test |
| Kelly et al. 2021 | low | low | low | low | low | low | low |
| Messner et al. 2001 | low | low | low | high | high | low | low |
| Oruc et al. 2021 | high | low | low | low | high | low | low |
| Shim et al. 2021 | high | low | low | low | high | low | low |
| Tanyeri Toker et al. 2023 | low | low | low | low | low | low | low |
| Tsuchiya et al. 2006 | high | low | low | high | low | low | low |

AABR = automated auditory brainstem response, TEOAE = transitory evoked otoacoustic emission.

### S1 File: Summary of findings table

**Population:** (Well) babies undergoing a two-stage hearing screening

**Setting:** Newborns from studies from all countries (high, middle and low-income countries) with the following criteria

1. Initial screening as an inpatient in the maternity clinic
2. Second test up to a maximum of one month later
3. No use of distortion product otoacoustic emissions (DPOAE)
4. Not exclusively NICU (neonatal intensive care unit).

Includes studies of well babies only and studies of well babies and NICU infants. Studies of exclusively NICU infants were not included in this review.

**Intervention:** Two-stage hearing screening using TEOAE, AABR, or combination of both

**Comparison:** not applicable

| Test findings |  |  | Certainty of the evidence (GRADE) | Comments |
| --- | --- | --- | --- | --- |
| Screening algorithm | Pooled refer rate (95% CI) | Number of newborns (studies) |  |  |
| AABR-AABR | 1.3% (0.9, 1.8%) | 331526 (18) | Moderate<sup>1</sup> | Moderate heterogeneity, but larger number of studies |
| TEOAE-TEOAE | 2.7% (2.2, 3.2%) | 671721 (55) | Moderate<sup>1</sup> | Moderate heterogeneity, but large number of studies |
| TEOAE-AABR | 3.9% (2.9, 5.1%) | 120054 (9) | Moderate<sup>2</sup> | Moderate heterogeneity and small number of studies |
| AABR-TEOAE | 5.9% (5.0, 6.9%) | 2316 (3) | Moderate<sup>2</sup> | No evidence of heterogeneity, but small number of studies |

CrI = Credibility interval, CI = confidence interval, GRADE = GRADE Working Group grades of evidence (high, moderate, low, very low).^1^Level of evidence was lowered by one level due to found moderate heterogeneity in random-effects meta-analysis. ^2^Level of evidence was lowered by one level due to small number of pooled studies in random-effects meta-analysis.

